# Specialty Choice Attitudes Among Medical Interns: Evidence from Hormozgan University of Medical Sciences

**DOI:** 10.64898/2026.06.12.26355502

**Authors:** Pourya Kashefi Sis, Ali Shendabadi, Nozhan Alimi, Elham Boushehri

## Abstract

**Background:** Choosing a medical specialty is a critical career decision that affects both physicians’ future professional lives and the composition of the healthcare workforce. Specialty preferences are shaped by multiple personal, educational, and socioeconomic factors, yet evidence from senior medical students in southern Iran remains limited. This study aimed to assess willingness to pursue specialty training among medical interns at Hormozgan University of Medical Sciences, identify their preferred specialties, and examine factors associated with their decisions.

**Methods:** This descriptive-analytical cross-sectional study was conducted in 2023 among medical interns at Hormozgan University of Medical Sciences in Bandar Abbas, Iran. Using a convenience census approach, all eligible interns were invited to participate, and 83 students completed an online questionnaire. The instrument collected demographic, academic, and occupational data, as well as reasons for willingness or unwillingness to pursue specialty training and specialty preferences. Content and face validity were assessed by faculty members and students, and internal consistency reliability in the present study was acceptable (Cronbach’s alpha = 0.82). Data were analyzed using descriptive statistics and logistic regression in SPSS version 27.

**Results:** Of the 83 participants, 50 (60.2%) reported willingness to pursue specialty training, while 33 (39.8%) did not. Among students willing to continue, the most frequently cited reasons were achieving a better economic position, broader job opportunities, and higher social status. Among those unwilling to continue, the most common reasons were fatigue from prolonged studying, financial problems, and the desire to start working after graduation. Radiology was the most common first-choice specialty, followed by otorhinolaryngology, dermatology, and cardiology. In regression analyses, no demographic or academic variable remained independently associated with willingness to pursue specialty training in the final multivariable model.

**Conclusions:** A majority of medical interns were interested in pursuing specialty training, with preferences concentrated in a limited number of specialties perceived as offering favorable financial prospects, prestige, and lifestyle. Economic concerns and educational fatigue were the dominant factors influencing willingness and unwillingness to continue specialty education. These findings highlight the need for structured career counseling, broader exposure to different specialties, and policy measures to address financial and structural barriers to residency training.

## Introduction

Choosing a medical specialty is one of the most important career decisions in medical training. This decision affects not only the future professional life of individual physicians, but also the composition and distribution of the healthcare workforce. Specialty preferences of medical students ultimately shape the future availability of specialists across disciplines, and imbalances in these preferences may contribute to shortages or oversupply in specific fields. For this reason, understanding how and why students develop interest in particular specialties is important for both educational planning and health workforce policy [1, 2].

Specialty choice is a complex and multifactorial process. Previous studies have shown that students’ decisions are influenced by a combination of personal interest, perceived aptitude, expected income, social prestige, lifestyle considerations, working conditions, duration of training, and exposure to positive or negative role models [3–8]. Clinical experiences appear to be particularly important, as students often modify their early preferences after entering the clinical environment and becoming familiar with the realities of different specialties [9, 10]. The internship period may therefore represent a particularly informative stage for assessing specialty intentions, because students have already experienced substantial clinical exposure and are approaching actual career decision-making.

Over the past decades, patterns of specialty preference have changed in many settings. Earlier studies often reported strong interest in major clinical fields such as surgery and internal medicine [11], whereas more recent evidence suggests increasing attraction toward specialties perceived to offer more controllable lifestyles, more favorable working hours, and better balance between professional and personal life [12]. At the same time, financial expectations and future job opportunities remain important determinants of career choice [1]. These trends have important implications for healthcare systems, especially where less preferred specialties may face workforce shortages [13].

In Iran, specialty selection is also shaped by the structure of medical education and the competitive national residency entrance process [14]. Although all medical graduates are potentially eligible to compete for residency training, access to preferred specialties depends heavily on examination performance and available training positions [15, 16]. As a result, some graduates may ultimately enter specialties that do not fully match their initial interests. Moreover, because exposure to all specialties during undergraduate training is not always uniform, students may make decisions based on incomplete experience or limited information. These issues make it especially important to examine students’ preferences and the factors influencing them in the local educational context.

Despite the practical importance of this topic, relatively few studies in Iran have focused specifically on senior medical students’ willingness to pursue specialty training and their specialty preferences, and available evidence has often been based on residents or graduates rather than students at the end of undergraduate medical education. Although we are aware of a few valuable recent research works [17–20]. In addition, local data from southern Iran and Bandar Abbas are limited. Given the potential influence of regional, cultural, educational, and labor-market factors on specialty choice, locally generated evidence is needed to better inform academic counseling and workforce planning.

Therefore, the present study was conducted to assess willingness to pursue specialty training among medical interns at Hormozgan University of Medical Sciences in Bandar Abbas, to identify their preferred specialties, and to examine the factors associated with their interest or lack of interest in continuing specialty education.

## Methods & Materials

### Study design and setting

This descriptive-analytical cross-sectional study was conducted in 2023 among medical interns enrolled at Hormozgan University of Medical Sciences, Bandar Abbas, Iran. The study aimed to assess students’ willingness or unwillingness to pursue specialty training, their specialty preferences, and the factors influencing these decisions.

### Data

The study population consisted of all medical students in the internship stage who were studying at Hormozgan University of Medical Sciences in 2023. Sampling was performed using a convenience census approach. After obtaining the necessary administrative approvals and ethics approval, all eligible interns were invited to participate in the study. We gathered data during the period of September to November 2023. Based on the Morgan table for sample size determination in descriptive studies, and considering that the total number of medical interns was 129, the minimum required sample size was estimated at 75 students. To compensate for potential non-response, 10% was added, and the target sample size was set at 83 participants. Ultimately, 83 students completed the questionnaire and were included in the analysis. The inclusion criteria were: being a medical student at Hormozgan University of Medical Sciences in the internship stage in 2023 and providing informed consent to participate in the study. The exclusion criterion was refusal to participate in the study.

Data were collected using an online questionnaire administered through Google Forms. The questionnaire was based on the instrument developed by Hashemipour et al [21]. for assessing factors affecting students’ willingness to continue education, for which prior psychometric evaluation had been reported. In the original study, internal consistency reliability was reported with a Cronbach’s alpha of 0.87. The questionnaire used in the present study consisted of several sections, including: demographic and personal information, items related to reasons for interest in continuing specialty education (8 items), items related to reasons for unwillingness to continue specialty education (9 items) and items related to reasons for choosing a particular specialty (5 items). The questionnaire also included items on specialty preferences and background characteristics such as age, sex, marital status, admission type/quota, national entrance examination rank, internship duration, employment status, personal income, interest in medicine, and grade point average. The motivation-related items were designed on a three-point scale consisting of important, relatively important, and not important. The questionnaire link was electronically distributed to students individually through virtual communication channels.

To assess content validity, the questionnaire was reviewed by university specialists and faculty members, who evaluated the relevance of the items to the study objectives and the adequacy of coverage of the main dimensions of the topic. Face validity was assessed by providing the questionnaire to 10 medical students, who reviewed the items in terms of clarity, simplicity, and comprehensibility. Their feedback was used to make final revisions to the questionnaire. To evaluate reliability in the present study, the questionnaire was completed by 10 students, and internal consistency was assessed using Cronbach’s alpha. The resulting Cronbach’s alpha coefficient was 0.82, indicating acceptable reliability of the instrument for measuring the study variables.

After approval of the study and receipt of the ethics code, the online questionnaire was prepared and distributed to eligible medical interns. Participation was voluntary. Before completing the questionnaire, students were informed about the purpose of the study, and only those who agreed to participate completed the form. Responses were collected electronically and then entered into SPSS version 27 for statistical analysis. The collected dataset is also accessible on the Harvard Dataverse platform [22].

The primary outcome variable was willingness to pursue specialty training, which was recorded as a binary variable (yes/no). Other variables included:

- demographic variables: age, sex, and marital status;
- academic variables: national entrance examination rank, internship duration, and GPA;
- admission-related variables: admission type/quota;
- occupational/economic variables: part-time job status;
- motivational variables: reasons for willingness or unwillingness to continue specialty training; and
- preferred specialty choices, recorded as first, second, and third priorities.

## Statistical analysis

Data were analyzed using SPSS software version 27 (IBM Corp., Armonk, NY, USA). Descriptive statistics were used to summarize the data. Quantitative variables were reported as mean ± standard deviation and range, while qualitative variables were presented as frequency and percentage.

To evaluate factors associated with willingness to pursue specialty training, logistic regression analysis was performed. First, univariable logistic regression models were fitted. Then, adjusted models controlling for age and sex were examined. Finally, a multiple logistic regression model was constructed including variables of interest and/or variables that showed significance in earlier analyses. Odds ratios (ORs) with 95% confidence intervals (CIs) were reported. A two-sided P-value of less than 0.05 was considered statistically significant.

## Ethical considerations

All ethical principles of research involving human participants were observed. The study was approved by the Ethics Committee of Hormozgan University of Medical Sciences under ethics code IR.HUMS.REC.1402.130 on July 8, 2023. Participation was voluntary, and informed consent was obtained from all participants before data collection. Confidentiality of participants’ information was maintained throughout the study.

## Results

### Participant characteristics

A total of 83 internship-stage medical students at Hormozgan University of Medical Sciences participated in this descriptive-analytical cross-sectional study in 2023 and completed the study questionnaire. The mean age of the participants was 26.69 ± 1.99 years (range: 23–40 years). Of the total sample, 39 students (47.0%) were female and 44 (53.0%) were male. Most participants were single (n = 65, 78.3%), including 27 women and 38 men. In addition, 11 students reported having a part-time job. A summary of the general demographic and academic characteristics of the participants are shown in table 2.

**Table 1.** Measurement characteristics of study variables.

| Scale | Measurement unit / method | Variable type |  |  | Role of variable |  |  | Variable title | No. |
| --- | --- | --- | --- | --- | --- | --- | --- | --- | --- |
|  |  | Qualitative | Discrete | Continuous | Control | Independent | Dependent |  |  |
| Ratio | Year |  |  | ✓ |  |  | ✓ | Age | 1 |
| Nominal | Female / Male | ✓ |  |  |  |  | ✓ | Gender | 2 |
| Nominal | Single / Married | ✓ |  |  |  |  | ✓ | Marital status | 3 |
| Nominal | Unemployed / Employed | ✓ |  |  |  |  | ✓ | Student employment status | 4 |
| Nominal | Regional quota / Other | ✓ |  |  |  |  | ✓ | Type of admission quota | 5 |
| Ratio | Numeric categorization |  |  | ✓ |  |  | ✓ | University entrance exam rank | 6 |
| Ratio | Number of full months elapsed since the first day of internship |  | ✓ |  |  |  | ✓ | Months elapsed since the start of internship | 7 |
| Ratio | Mean score obtained for each questionnaire item |  |  | ✓ |  | ✓ |  | Score from the questionnaire on interest or lack of interest in continuing education | 8 |

**Table 2.** Demographic and Educational Findings of the Study.

| Variable | Number | Percentage (%) |
| --- | --- | --- |
| **Gender** |  |  |
| Male | 44 | 53.0 |
| Female | 39 | 47.0 |
| **Marital Status** |  |  |
| Married | 18 | 21.7 |
| Single | 65 | 78.3 |
| **Part-time Job** |  |  |
| No | 72 | 86.7 |
| Yes | 11 | 13.3 |
| **National University Entrance Exam Rank** |  |  |
| Below 1000 | 4 | 4.8 |
| 1000 to 2000 | 17 | 20.5 |
| 2000 to 4000 | 36 | 43.4 |
| Above 4000 | 26 | 31.3 |
| **Admission Type** |  |  |
| Type 1 | 6 | 7.2 |
| Type 2 | 35 | 42.2 |
| Type 3 | 22 | 26.5 |
| Other | 1 | 1.2 |
| Deprived areas quota | 11 | 13.3 |
| Veterans/Sacrificers quota | 8 | 9.6 |
| **Months Elapsed Since Internship Start** |  |  |
| 1 | 1 | 1.2 |
| 2 | 1 | 1.2 |
| 3 | 5 | 6.0 |
| 6 | 2 | 2.4 |
| 7 | 2 | 2.4 |
| 8 | 1 | 1.2 |
| 9 | 6 | 7.2 |
| 10 | 2 | 2.4 |
| 12 | 12 | 14.5 |
| 13 | 13 | 15.7 |
| 14 | 11 | 13.3 |
| 15 | 13 | 15.7 |
| 16 | 10 | 12.0 |
| 17 | 1 | 1.2 |
| 18 | 3 | 3.6 |
| **Medical School GPA** |  |  |
| 14–15 | 6 | 7.2 |
| 15–16 | 31 | 37.3 |
| 16–17 | 33 | 39.8 |
| 17–18 | 8 | 9.6 |
| 18–19 | 4 | 4.8 |
| 19–20 | 1 | 1.2 |
| **Total** | **83** | **100.0** |

National entrance examination rank was categorized into four levels; the most frequent category was a rank between 2,000 and 4,000 (n = 36), while the least frequent was rank below 1,000 (n = 4). Regarding admission quota type, Region 2 quota was the most common (n = 35), whereas “other” admission type had the lowest frequency (n = 1). The number of months completed in internship was distributed across a relatively wide range (1–18 months), with the highest frequencies observed at months 13 and 15 (n = 13 each), and the lowest frequencies at months 1, 2, 8, and 17 (n = 1 each). For grade point average (GPA) during medical training, the 16–17 category was the most common (n = 33), whereas the 19–20 category was the least frequent (n = 1).

Among the 83 participating medical interns, 50 students (60.2%) reported willingness to continue their education in a specialty training program, while 33 students (39.8%) stated that they were not willing to pursue specialty education.

### Reasons for willingness or uwillingness to pursue specialty training

The main motivations reported by students who expressed willingness to enter specialty training are summarized in table 3. The most frequently cited reason was achieving a better economic position (n = 41), making it the leading motivation for pursuing specialty education. This was followed by access to better and broader job opportunities (n = 35) and obtaining a higher social status (n = 34). These findings indicate that economic and social advancement were the dominant drivers of interest in specialty training.

**Table 3.**
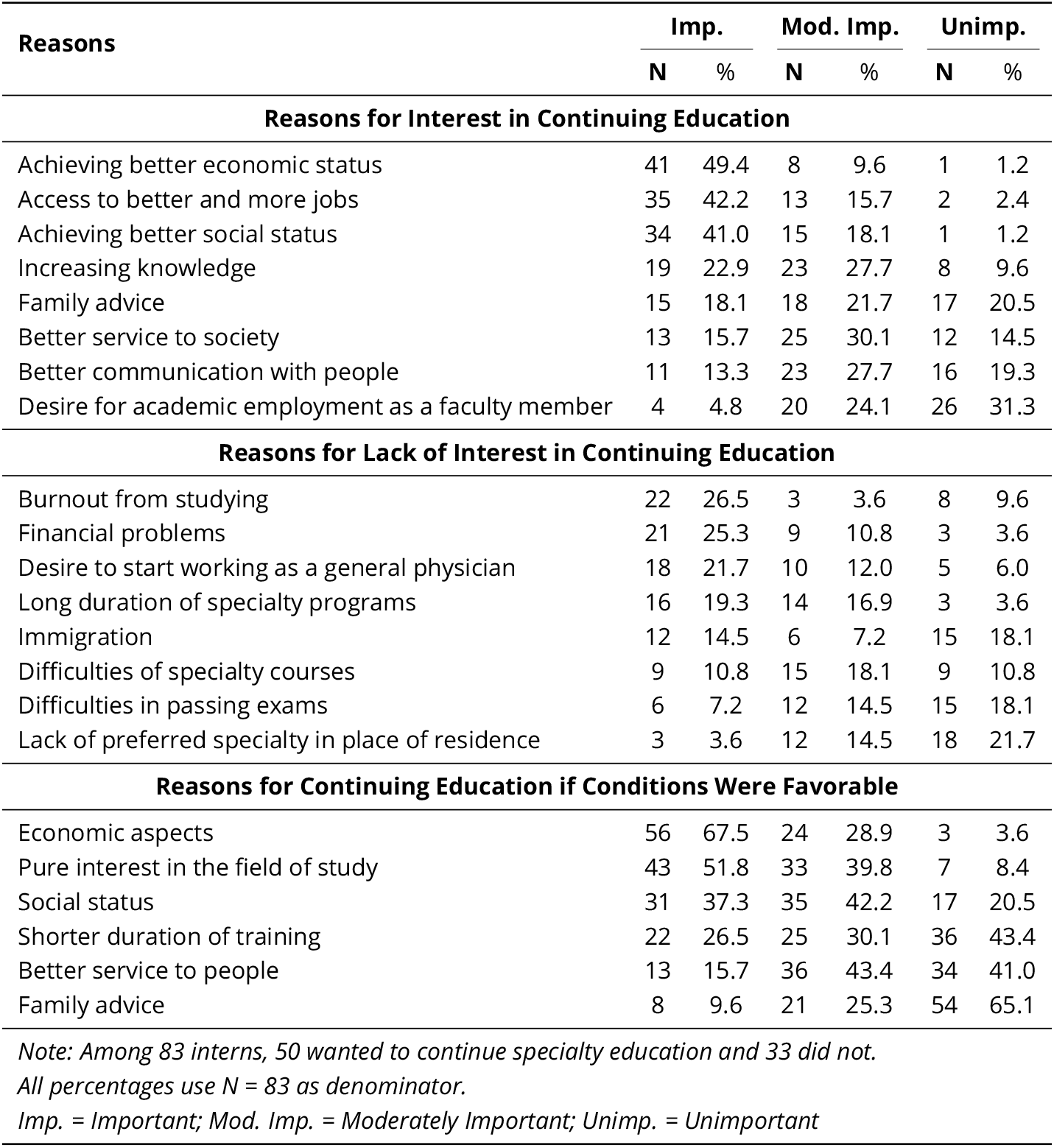
Interns’ Perspectives on Continuing Education.

Other reasons were reported less frequently. Increasing knowledge was mentioned by 19 students, and family recommendation was reported by 15 students. Better service to society and improved communication with people were selected by 13 and 11 students, respectively. The least frequently reported motivation was the desire to be employed as an academic faculty member at a university, which was mentioned by only 4 students. Overall, the pattern suggests that personal and material incentives were more prominent than academic or altruistic considerations.

Among the 33 students who were not willing to continue specialty training, the reasons for this decision are presented in table 3. The most common reason was fatigue from prolonged studying, reported by 22 students, indicating the importance of educational burnout in shaping students’ decisions. Financial problems were the second most frequently reported factor (n = 21), underscoring the role of economic constraints in discouraging continued education.

The third most common reason was the desire to start working after graduation (n = 18), suggesting that a substantial proportion of students preferred entering the workforce immediately after completion of general medical training. This was followed by the long duration of specialty training programs (n = 16) and plans to immigrate (n = 12). Fewer students cited the difficulty of specialty courses (n = 9) and problems succeeding in examinations (n = 6). The least frequently reported reason was the unavailability of the preferred specialty in the place of residence (n = 3). In general, individual and psychological factors—particularly study fatigue—along with financial concerns appeared to play a greater role than structural or geographic factors in explaining unwillingness to pursue specialty education.

Students were also asked which factors would influence their decision to pursue specialty education under more favorable circumstances. For students who were currently unwilling to continue specialty training, this meant imagining that the barrier underlying their unwillingness had been removed and then reporting what factors would influence the specialty they would choose. For those who had already expressed willingness to continue, it referred to the factors that, if circumstances were more favorable, could shape their choice of a different specialty field. As shown in table 3, the most frequently cited factor was economic benefit, reported by 56 students (67.5%). This finding highlights the central role of financial return and professional prospects in students’ decision-making.

The second most common factor was pure interest in the scientific content of the specialty, mentioned by 43 students (51.8%). Social position and prestige ranked third, reported by 31 students (37.3%). A shorter duration of training was cited by 22 students (26.5%), suggesting that the length of specialty education remains an important deterrent. Better service to people was reported by 13 students (15.7%), and family advice was the least cited factor, mentioned by 8 students (9.6%). Taken together, these findings suggest that under more favorable circumstances, students’ decisions to pursue specialty training would still be driven primarily by economic and scientific motivations rather than family influence or altruistic considerations.

### Specialty preferences

The distribution of students’ first, second, and third specialty choices is shown in figure 1. Analysis of first-priority choices demonstrated that radiology was the most preferred specialty, with 24 students selecting it as their first choice. Otorhinolaryngology (ENT) ranked second with 18 first-priority selections, followed by dermatology with 9 selections. Other specialties receiving first-priority selections included cardiology (n = 7), psychiatry (n = 4), and ophthalmology (n = 3). In contrast, several specialties—including emergency medicine, neurology, anesthesiology, internal medicine, social medicine, and sports medicine—received no first-priority selections.

**Figure 1.**
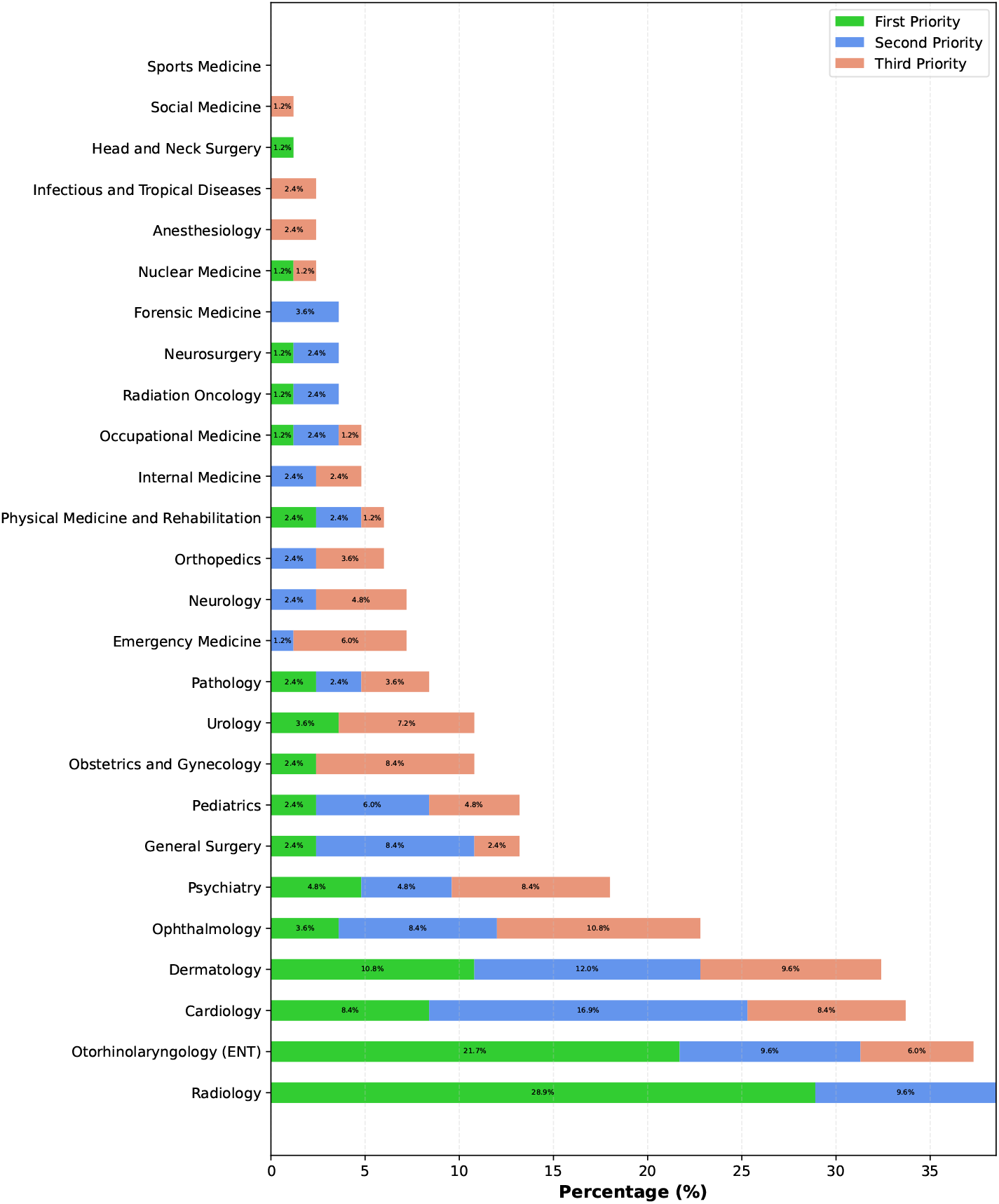
Priorities of Hormozgan University of Medical Sciences interns for medical specialty selection. This chart illustrates the distribution of first, second, and third preferences across 26 specialties. Data are presented as percentages (N=83 total respondents). Radiology shows the highest proportion of first-priority selections (28.9%), followed by Otorhinolaryngology (21.7%) and Dermatology (10.8%).

For second-priority choices, cardiology ranked first with 14 selections, indicating that although it was not among the most frequently chosen first priorities, it remained an important option for many students. Dermatology was the second most common second-priority choice (n = 10), followed by radiology and ENT (n = 8 each). Some specialties, including obstetrics and gynecology, urology, anesthesiology, infectious diseases, social medicine, and sports medicine, received no second-priority selections. The pattern changed somewhat for third-priority preferences. Ophthalmology had the highest number of third-priority selections (n = 9), followed by dermatology (n = 8). Cardiology, psychiatry, and obstetrics and gynecology each received 7 third-priority selections. Meanwhile, radiology, radiation oncology, neurosurgery, forensic medicine, head and neck surgery, and sports medicine received no third-priority selections. Overall, the data indicate a strong preference for specialties such as radiology, ENT, dermatology, and cardiology. These specialties may be perceived by students as offering more favorable career prospects, social prestige, or work-life balance. In contrast, less preferred specialties either appeared only in lower-priority rankings or were not selected at all.

The results of simple, age- and sex-adjusted, and multiple logistic regression analyses examining factors associated with willingness to choose a specialty are presented in table 4. In the final multiple logistic regression model, none of the studied variables remained statistically significant. Although in the adjusted model (controlling for age and sex), total GPA in the 16–17 range (P = 0.049) and 17–18 range (P = 0.028) reached statistical significance, these associations were not confirmed in the final multivariable model. In the final model, the corresponding P-values increased to 0.439 and 0.130, respectively, and the 95% confidence intervals for the odds ratios included 1.

**Table 4.** Factors Influencing the Tendency to Choose a Specialty.

| Variable | Simple Logistic Regression (Univariate) |  | Logistic Regression Adjusted for Age and Gender |  | Multiple Logistic Regression (Final Model) |  |
| --- | --- | --- | --- | --- | --- | --- |
|  | P value | OR (95% CI) | P value | OR (95% CI) | P value | OR (95% CI) |
| Age (per 1 year) | 0.792 | 1.0 (0.8 – 1.3) | - | - | 0.721 | 0.9 (0.7 – 1.2) |
| Gender (Male) | 0.262 | 1.7 (0.7 – 4.0) | - | - | 0.221 | 1.8 (0.7 – 4.6) |
| Single | 0.246 | 0.5 (0.2 – 1.5) | 0.412 | 0.6 (0.3 – 1.2) | - | - |
| Part-time job (Yes) | 0.134 | 0.3 (0.1 – 1.4) | 0.175 | 0.4 (0.1 – 2.1) | - | - |
| Admission quota (Region 1) | - | 1 | - | 1 | - | - |
| Admission quota (Region 2) | 0.758 | 1.3 (0.2 – 8.1) | 0.999 | 0.1 (0.1 – 1.4) | - | - |
| Admission quota (Region 3) | 0.365 | 2.4 (0.3 – 15.6) | 0.412 | 2.2 (0.8 – 7.9) | - | - |
| Admission quota (Other region) | 0.812 | 0.1 (0.1 – 1.0) | 0.999 | 0.1 (0.1 – 1.1) | - | - |
| Admission quota (Deprived areas) | 0.901 | 1.1 (0.2 – 9.7) | 0.177 | 4.2 (0.6 – 25.4) | - | - |
| Admission quota (Veterans/Sacrificers) | 0.363 | 0.3 (0.1 – 4.4) | 0.808 | 0.6 (0.1 – 2.6) | - | - |
| National rank (Below 1000) | - | 1 | - | 1 | - | - |
| National rank (1000 to 2000) | 0.749 | 0.7 (0.1 – 6.2) | 0.646 | 0.4 (0.1 – 13.2) | - | - |
| National rank (2000 to 4000) | 0.916 | 0.9 (0.1 – 7.0) | 0.486 | 0.3 (0.1 – 8.4) | - | - |
| National rank (Above 4000) | 0.361 | 0.4 (0.1 – 3.1) | 0.432 | 0.2 (0.1 – 2.6) | - | - |
| Overall GPA (14–15) | - | 1 | - | 1 | - | - |
| Overall GPA (15–16) | 0.942 | 0.9 (0.2 – 5.4) | 0.182 | 0.1 (0.1 – 2.7) | - | - |
| Overall GPA (16–17) | 0.439 | 0.5 (0.1 – 2.2) | **0.049\*** | 0.1 (0.1 – 1.0) | 0.413 | 0.4 (0.2 – 1.8) |
| Overall GPA (17–18) | 0.148 | 0.1 (0.1 – 1.9) | **0.028\*** | 0.1 (0.1 – 0.6) | 0.130 | 0.2 (0.1 – 1.0) |
| Overall GPA (18–19) | 0.451 | 1.0 (0.6 – 12.2) | 0.190 | 0.1 (0.1 – 4.5) | - | - |
| Overall GPA (19–20) | 0.573 | 5.9 (0.8 – 27.4) | 0.999 | 6.8 (0.9 – 31.2) | - | - |

\* P value < 0.05

Other variables, including age, sex, marital status, having a part-time job, admission quota category, national entrance examination rank, and other GPA categories (14–15, 15–16, 18–19, and 19–20), were not significantly associated with willingness to pursue specialty training in any of the regression models (all P > 0.05). Specifically, age (OR = 0.90, P = 0.721) and male sex (OR = 1.80, P = 0.221) showed no significant relationship with specialty preference. Similarly, none of the admission quota categories (Region 1, Region 2, Region 3, deprived area quota, veteran quota, or other) had a statistically significant effect on willingness to choose a specialty. Overall, the regression analysis did not identify any independent demographic or academic predictors of willingness to pursue specialty training in this sample.

## Discussion

This study examined specialty preferences, willingness to pursue residency training, and the main motivating and discouraging factors among interns at Hormozgan University of Medical Sciences. Overall, the findings suggest that specialty choice is a multifactorial decision shaped primarily by economic considerations, perceived job opportunities, social prestige, and lifestyle-related expectations, rather than by basic demographic or academic characteristics alone. These findings are important because the specialty preferences of current medical students contribute directly to the future composition of the physician workforce, and imbalances in specialty distribution may ultimately affect health system performance and access to care [1, 23]. Specialty choice has long been recognized as a key stage in medical career development. A central finding of the present study was that a majority of students expressed interest in pursuing specialty training. This is generally consistent with reports from many settings indicating that interest in specialization has increased over time [5].

In the present study, students’ preferences were concentrated in specialties such as radiology, otorhinolaryngology, dermatology, and cardiology, whereas several other specialties attracted little or no interest. This pattern is noteworthy because it appears to differ from the traditional dominance of major or highly demanding specialties reported in older literature [11, 12]. The current findings instead suggest a shift toward specialties perceived as offering more favorable working conditions, more controllable lifestyles, and attractive financial prospects [14]. This pattern is in line with more recent international evidence showing that younger physicians increasingly value work-life balance alongside income and prestige [24]. In this framework, specialties such as dermatology, ophthalmology, and radiology have become especially attractive because they are often associated with more predictable schedules, less physically demanding workloads, and comparatively favorable remuneration. This shift away from some major specialties may reflect several realities experienced by students during clinical training. Prolonged exposure to demanding hospital environments, heavy workloads, high emotional burden, difficult interpersonal climates in some departments, and concerns about unclear or stressful future career conditions may all reduce the attractiveness of more intensive specialties [7]. In addition, limited or uneven exposure to certain specialties during clinical rotations may influence students’ awareness of those fields and reduce the likelihood that they will consider them seriously [25]. Thus, specialty preference may be shaped not only by the inherent nature of a field, but also by the quality and extent of students’ contact with it during undergraduate training.

Economic considerations emerged as one of the strongest themes in this study, both among students interested in specialty training and among those who were hesitant or unwilling to continue. This finding is highly meaningful in the context of medical education, where students spend many years in training before reaching stable income. The long educational pathway in medicine can create pressure to choose specialties that promise faster financial return, greater earning potential, or more secure employment opportunities. Conversely, when the perceived financial burden of continued training is high, motivation to pursue residency may decline. The prominence of financial factors in the present study is therefore consistent with previous research showing that income expectations, living conditions during residency, and anticipated future earnings are major determinants of specialty preference [1, 4, 26–30]. The importance of economic motives in this study should not be interpreted narrowly as simple materialism. Rather, it may reflect a broader concern with long-term professional stability, independence, and quality of life. Students often evaluate specialties not only in terms of scientific interest, but also in relation to the years required for training, the financial sacrifices expected during residency, and the sustainability of the eventual career path. This interpretation is supported by another finding of the present study: Under more favorable conditions, for both students who were willing and those who were unwilling to pursue specialty training, economic considerations were the most frequently reported motivation for continuing specialty education.

Another important finding was that fatigue from studying was one of the most common reasons for not pursuing specialty training. This result highlights the role of academic burnout in medical career decision-making. The prolonged duration of medical education, repeated high-stakes evaluations, and cumulative psychological and physical demands of the clinical years may contribute to exhaustion by the end of internship. For some students, the prospect of entering another long and demanding period of residency training may therefore be discouraging, even if they remain interested in medicine as a profession. Similarly, the preference of some students to enter the job market immediately after graduation may represent both economic necessity and a desire to regain control over time, income, and personal life after years of intensive education. The findings also support the growing importance of lifestyle considerations in specialty choice. Although the questionnaire did not directly measure “controllable lifestyle” as a separate variable, the pattern of preferred specialties and the reasons reported by students strongly suggest that this concept is highly relevant. Recent studies in different countries have likewise shown that younger generations of medical trainees increasingly prioritize manageable work hours, reduced on-call burden, and the possibility of balancing professional and personal responsibilities [30, 31] and some research show rising regret rates in medical residents [32–34]. This trend may help explain declining interest in more demanding or stressful specialties and growing interest in fields traditionally associated with more regular schedules.

Immigration was an important reason for unwillingness to pursue specialty training in a notable minority of interns in our study. While less common than fatigue and financial concerns, this finding is broadly consistent with a recent study [35] among Iranian residents reporting high immigration willingness, and may indicate that immigration-related thinking begins to affect career decisions even before residency entry. In the context of the broader and ongoing brain drain in Iran, this finding warrants particular attention from policymakers. Efforts to strengthen the medical workforce should therefore address conditions that may encourage young physicians to consider leaving the country, thereby increasing the risk of future medical workforce loss.

In the present study, no statistically significant independent association was found between willingness to pursue specialty training and demographic or academic variables such as age, sex, entrance examination rank, admission quota, or GPA in the final regression model. This finding reinforces the idea that specialty choice is a complex process not easily explained by a limited set of measurable background variables. Although GPA categories showed temporary significance in the age- and sex-adjusted model, this relationship did not persist in the final multivariable analysis. The absence of stable predictors suggests that unmeasured factors—such as personality traits [36, 37], lifestyle values [38], role models [39, 40], prior clinical experiences [9, 10], perceptions of residency culture, and broader cultural expectations—may play a more important role.

The lack of a significant gender difference in the present study is also notable. In many previous studies, gender has been reported as an important factor in specialty choice, often with female students showing greater preference for specialties compatible with family life and less preference for highly demanding or time-intensive fields [5, 15, 24, 41]. In our study, however, such a difference was not statistically evident. This may indicate changing patterns of expectation among both male and female students, or it may reflect the limited sample size and local context. Therefore, although gender did not emerge as an independent predictor here, it should not be dismissed as irrelevant without further multicenter and larger-scale investigation.

Similarly, marital status was not identified as an independent predictor in the final model, but it remains conceptually important. Family responsibilities, emotional support, financial stability, and future household planning may all influence decisions about whether and when to enter specialty training. In this sense, the absence of a statistical association in the present sample should be interpreted cautiously. Such factors may operate indirectly or interact with variables not captured in this study [8, 24].

The relatively lower importance of family recommendation compared with financial and personal motives is another meaningful finding. It suggests that, in this group of students, specialty selection was driven more by individual career planning than by direct family pressure. Nevertheless, the broader literature indicates that family background may still matter, especially when students have physician parents or close relatives who can serve as role models, provide information about certain specialties, or create practical opportunities for future professional entry [29, 42]. Therefore, family influence may be subtle and indirect rather than explicitly stated by students.

These findings also draw attention to the possible role of educational counseling and career guidance. In many medical schools, students receive insufficient structured information about the full range of available specialties, the realities of residency training, and long-term career trajectories [1]. In such situations, students may rely heavily on informal student culture, departmental atmosphere, anecdotal impressions, or short-term experiences during rotations [25]. This may lead to exaggerated enthusiasm for some specialties and unfair discouragement from others. A lack of formal advising may also result in mismatch between a student’s personality, values, capabilities, and eventual specialty choice. Individual or group-based career counseling, mentorship from faculty, and structured orientation sessions about residency options may therefore improve the quality of decision-making and lead to better alignment between student preferences and health system needs [43].

From a policy perspective, the findings have several implications. First, the concentration of interest in a limited number of specialties and the low attractiveness of several others may contribute over time to imbalances in the physician workforce. If such patterns continue, some fields may face oversupply while others experience persistent shortages [13]. Second, because economic concerns were strongly linked to both willingness and unwillingness to continue specialty training, policymakers should consider improving financial support mechanisms, residency conditions, and future career incentives in less popular specialties. Third, because clinical exposure appears to shape preferences substantially, improving the educational environment of clinical departments may influence how students perceive different specialties.

This study has several strengths. One important strength is the high response rate, which may have been facilitated by careful electronic distribution of the questionnaire to students on an individual basis. Although this method was time-consuming, it appears to have improved participation and reduced missing data. Another strength is the inclusion of students in the internship stage, which is an especially relevant period for assessing specialty intention because students have already experienced substantial clinical exposure and are closer to making actual residency decisions.

At the same time, several limitations should be acknowledged. First, the cross-sectional design does not allow causal inference and cannot capture how preferences may change over time. Specialty interest is dynamic, and a longitudinal design would provide a clearer understanding of how clinical experiences, life events, and educational progression influence decision-making [44–46]. Second, the sample size was relatively small, which may have limited statistical power, particularly in regression analyses and subgroup comparisons. Third, the study was conducted in a single university, and therefore the findings may not be fully generalizable to students in other parts of Iran or in different educational settings. Multicenter studies with larger and more diverse samples are needed to examine whether the observed pattern is specific to this setting or reflects a broader trend.

Future research should therefore move in several directions. Studies involving multiple universities, including both public and private institutions, would improve generalizability. Longitudinal studies following students from pre-clinical years through internship and into residency application would be especially valuable for documenting changes in specialty preference over time. It would also be useful to examine factors not directly assessed in the present study, such as personality dimensions, perceived departmental climate, exposure to role models, and family medical background.

## Conclusion

In conclusion, this study provides a useful picture of specialty preferences and their determinants among medical interns in Hormozgan University of Medical Sciences. The findings suggest that students are increasingly drawn to specialties perceived as offering better financial prospects, more desirable social status, and a more manageable lifestyle, while interest in more demanding specialties appears to be lower. At the same time, no independent demographic or academic predictor was identified in the final regression model, underscoring the complexity of specialty choice. These results highlight the importance of targeted career counseling, improved educational exposure to different specialties, and policy interventions aimed at reducing financial and structural barriers to residency training. Such efforts may help align student preferences more effectively with both personal career satisfaction and the long-term needs of the health system.

## Data Availability

All data produced are available online at Harvard Dataverse platform

https://dataverse.harvard.edu/dataset.xhtml?persistentId=doi:10.7910/DVN/7UMX92

## Acknowledgment

The authors used GPT-5.4 (OpenAI) to assist with language editing and improvement of grammar and readability. The authors reviewed and edited all output and take full responsibility for the final content of the manuscript.

